# Whole-genome sequencing study of 488,888 individuals identifies rare variants and genes associated with glaucoma

**DOI:** 10.1101/2025.06.02.25328820

**Authors:** Xiaoyi Raymond Gao, Krishnakumar Kizhatil

## Abstract

Glaucoma is a leading cause of irreversible blindness worldwide, with primary open-angle glaucoma (POAG) being the most common form. While previous genome-wide association studies have identified common genetic variants associated with POAG, primarily through array-based genotyping and imputation, the role of rare variants remains poorly understood. Our study aims to address this gap in knowledge by conducting the largest whole-genome sequencing (WGS) analysis of glaucoma to date. We analyzed WGS data from 488,888 UK Biobank participants, comprising 19,440 POAG cases and 469,448 controls. To validate our findings, we used the FinnGen cohort and the Million Veteran Program. We confirmed associations of known glaucoma genes, including *MYOC*, *OPTN*, and identified more than 68 previously unreported genes for glaucoma, such as *FMO4*, *PRRC2C*, *MTOR*, *ANGLPL7*, *AREL1*, and *LINC02210-CRHR1*. Seven of these genes, i.e., *MTOR*, *COL1A2*, *DDR2*, *LYN*, *NR1H3*, *TGFB3*, and *MAPT*, are current drug targets. Our findings underscore the value of WGS for uncovering rare variants and novel genes implicated in glaucoma. By expanding the genetic architecture of this complex disease, our work paves the way for future research into disease mechanisms, and ultimately, the development of more targeted preventive and therapeutic strategies.

## Introduction

Glaucoma remains the leading cause of irreversible blindness worldwide^1,2^, characterized by progressive optic nerve head excavation and irreversible visual field loss. Primary open-angle glaucoma (POAG), accounting for approximately 90% of glaucoma cases in North America, represents the most prevalent form of this condition^3,4^. POAG demonstrates high heritability, with numerous genetic loci identified through previous genome-wide association studies (GWASs)^5–7^.

While these earlier GWASs have significantly advanced our understanding of glaucoma genetics, they have predominantly relied on genome-wide arrays and imputation techniques. This approach has several inherent limitations. Array-based studies primarily capture common genetic variants, which typically exhibit modest effect sizes. Furthermore, imputation methods often fail to adequately represent rare variants, leaving substantial gaps in our understanding of the complete genetic architecture of POAG. Consequently, the contribution of rare variants to glaucoma pathogenesis remains largely unexplored, despite their potential for larger effect sizes and greater clinical significance.

The recent release of whole-genome sequencing (WGS) data from the UK Biobank (UKB)^8^, a prospective cohort of approximately half a million adult participants, presents an unprecedented opportunity to investigate rare genetic variants and to identify novel genes across numerous human traits and diseases^9,10^. Previous studies leveraging large-scale sequencing have successfully identified rare variants with substantial effect sizes that demonstrate considerable translational potential, such as *GHRH* variants for height determination and *PCSK9* variants as targets for low-density lipoprotein reduction^11,12^. Despite this potential, the UK Biobank’s WGS data remains largely unreported specifically for glaucoma genetics.

In this study, we analyzed WGS data from 488,888 UKB participants to identify rare variants and genes associated with POAG. We further validated our findings using summary statistics from the FinnGen cohort and the Department of Veterans Affairs (VA) Million Veteran Program (MVP). To our knowledge, this represents the largest sequencing study of glaucoma conducted to date. Our comprehensive approach has uncovered previously unreported rare variants and genes associated with glaucoma, substantially expanding our understanding of the biological mechanisms underlying this disease and revealing potential novel drug targets for therapeutic intervention.

## Results

In this study, we analyzed WGS data from 488,888 UKB participants, of whom 464,778 (95%) were of European (EUR) ancestry. The cohort had a mean age of 57.5 years (standard deviation: 8.4 years) with 54% female participants. We employed a comprehensive suite of state-of-the-art analytical approaches to identify variants, genes, potential biological mechanisms, and drug targets associated with POAG.

### Single-Variant Analysis

Our single-variant analysis identified 6,787 variants in 196 genes (excluding intergenic regions) significantly associated with POAG (*P* < 5×10^−8^, Supplementary Table 1). Among these, we discovered 56 previously unreported genes for POAG, with 42 identified in the EUR analysis and 14 additional genes in the pan-ancestry analysis (Table 1).

**Table 1.**
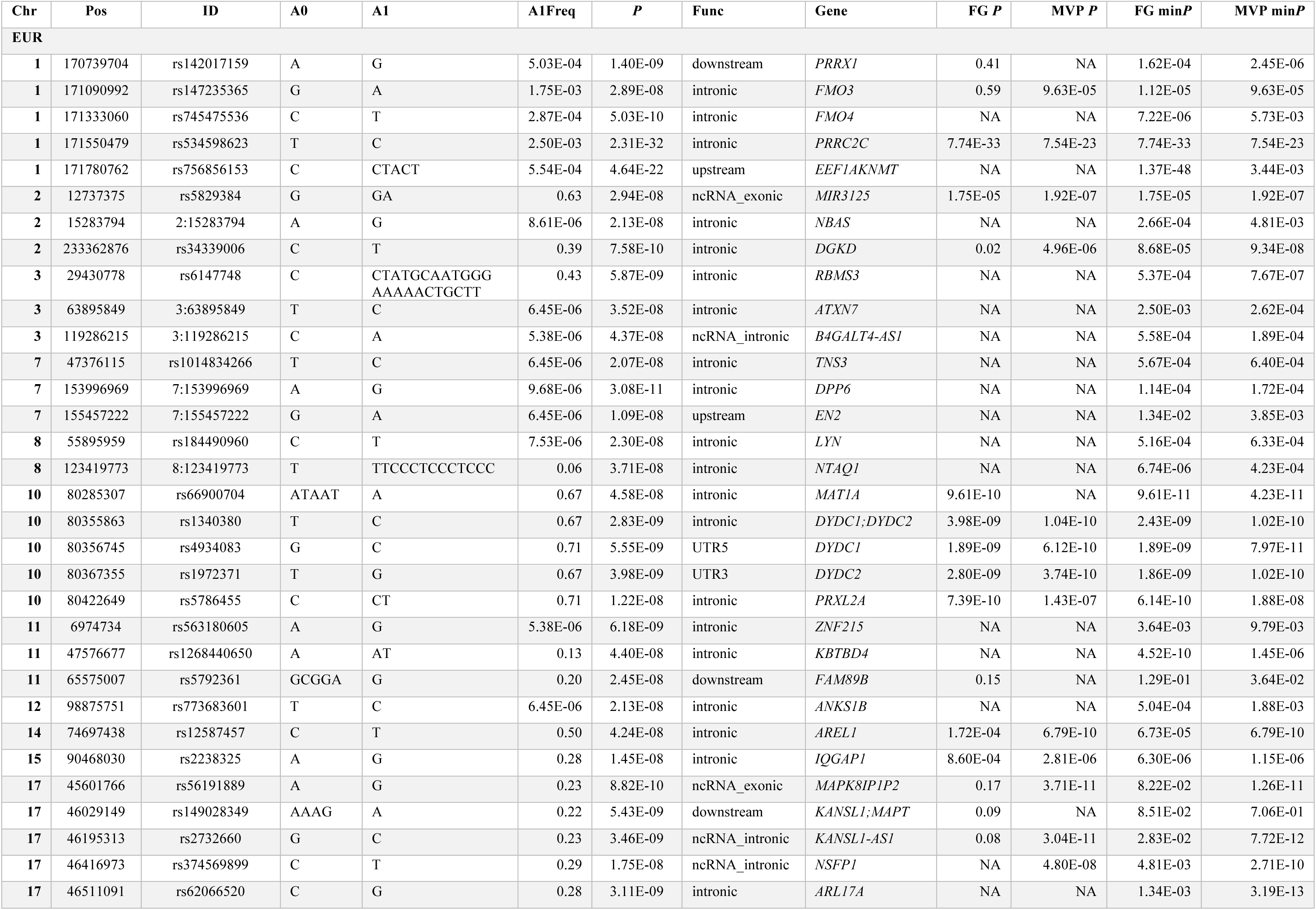
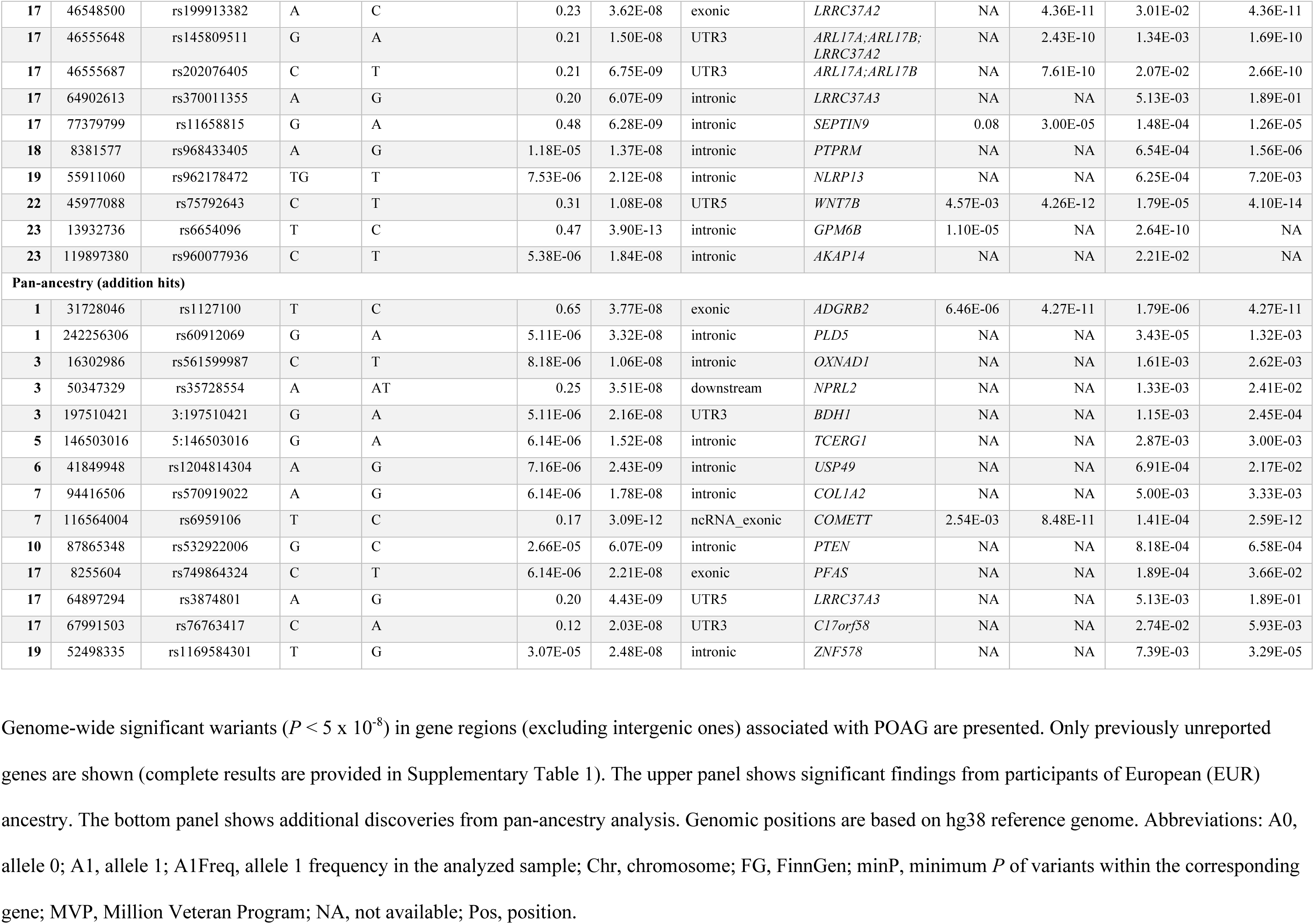
Genome-wide significant variants and their corresponding genes for primary open-angle glaucoma.

We confirmed significant associations with known glaucoma-related rare variants, including rs74315329 (allele frequency [AF] = 0.013%, *P* = 3.49×10^−50^) in *MYOC* and rs28939688 (AF = 0.0005%, *P* = 1.25×10^−8^) in *OPTN*. Additionally, we identified novel rare variants significantly associated with POAG, such as rs147235365 in *FMO3* (AF = 0.018%, odds ratio [OR] = 1.84, *P* = 2.89×10^−8^) and rs534598623 in *PRRC2C* (AF = 0.025%, OR = 2.70, *P* = 2.31×10^−32^). The *FMO3* variant was replicated in MVP (*P* = 9.63×10^−5^), while the *PRRC2C* variant was replicated in both FinnGen (*P* = 7.74×10^−33^) and MVP (*P* = 7.53×10^−23^). Three additional rare variants demonstrated significant POAG associations: rs142017159 in *PRRX1* (AF = 0.05%, OR = 2.98, *P* = 1.40×10^−9^), rs745475536 in *FMO4* (AF = 0.03%, OR = 3.96, *P* = 5.03×10^−10^), and rs756856153 in *EEF1AKNMT* (AF = 0.06%, OR = 4.50, *P* = 4.64×10^−22^). These findings were replicated in independent cohorts: *PRRX1* in MVP (minimum *P* of variants in the gene [minP] = 2.45×10^−6^), *FMO4* in FinnGen (minP = 7.22×10^−6^), and *EEF1AKNMT* in FinnGen (minP = 1.37×10^−48^). We also identified multiple common variants in previously unreported genes significantly associated with POAG, including rs5829384 (*MIR3125*), rs34339006 (*DGKD*), rs66900704 (*MAT1A*), rs4934083 (*DYDC1*), rs1972371 (*DYDC2*), rs5786455 (*PRXL2A*), rs12587457 (*AREL1*), rs56191889 (*MAPK8IP1P2*), rs11658815 (*SEPTIN9*), and rs75792643 (*WNT7B*). All these variants were replicated in either FinnGen or MVP or both (*P* < 5×10^−5^). Replicated variants showed consistent effect direction in replication datasets.

In the pan-ancestry analysis, we identified additional significant associations, including common variants rs1127100 in *ADGRB2* and rs6959106 in *COMETT* (both replicated in MVP, *P* < 5×10^−11^). We also found rare variants rs60912069 in *PLD5* (AF = 0.0005%, *P* = 3.32×10^−8^) and rs1169584301 in *ZNF578* (AF = 0.003%, *P* = 2.48×10^−8^), with both genes showing associations in FinnGen (minP = 3.43×10^−5^) and MVP (minP = 3.29×10^−5^), respectively. Figure 1(a) presents a circular Manhattan plot of the genome-wide P-values for pan-ancestry results. The genomic control lambda values for EUR and pan-ancestry analyses were well-controlled at 0.86 and 0.87, respectively.

**Figure 1.**
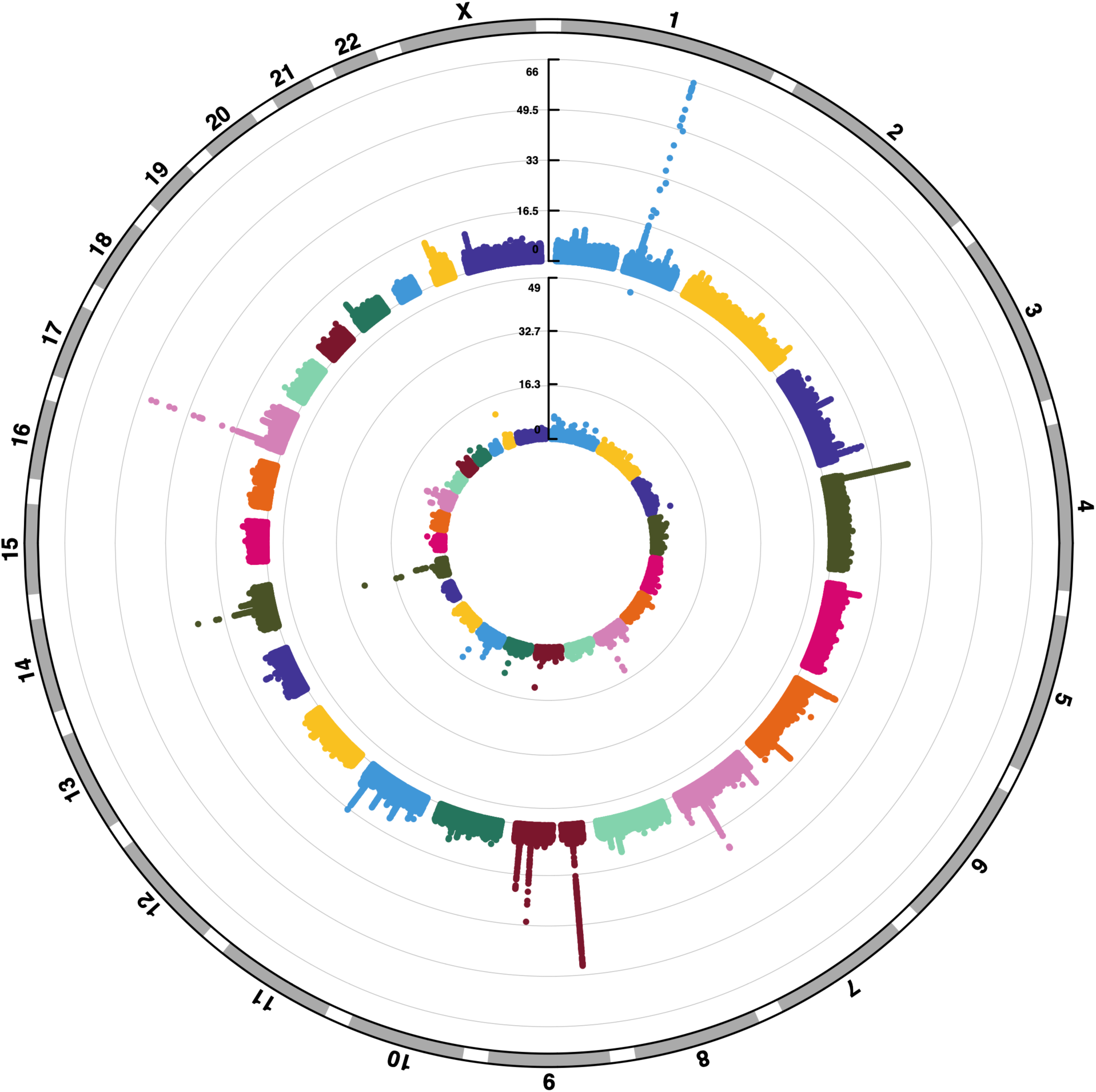
Manhattan plots displaying the -log_10_(*P*) for the association between POAG and single variants and genes. (a) Single-variant pan-ancestry results. (b) Gene-based pan-ancestry results. Genetic variants or genes are plotted by genomic position.

### Gene-Based Analysis of Rare Variants

Our REGENIE^13^ gene-based analysis of rare variants identified 47 genes significantly associated with POAG (*P* < 2.5×10^−6^), of which 12 were previously unreported discoveries. Specifically, 46 genes were identified from the EUR analysis with one additional gene from the pan-ancestry analysis; six genes emerged from the loss-of-function (LOF) analysis, while 41 were identified from the combined LOF and missense analysis (Table 2).

**Table 2.**
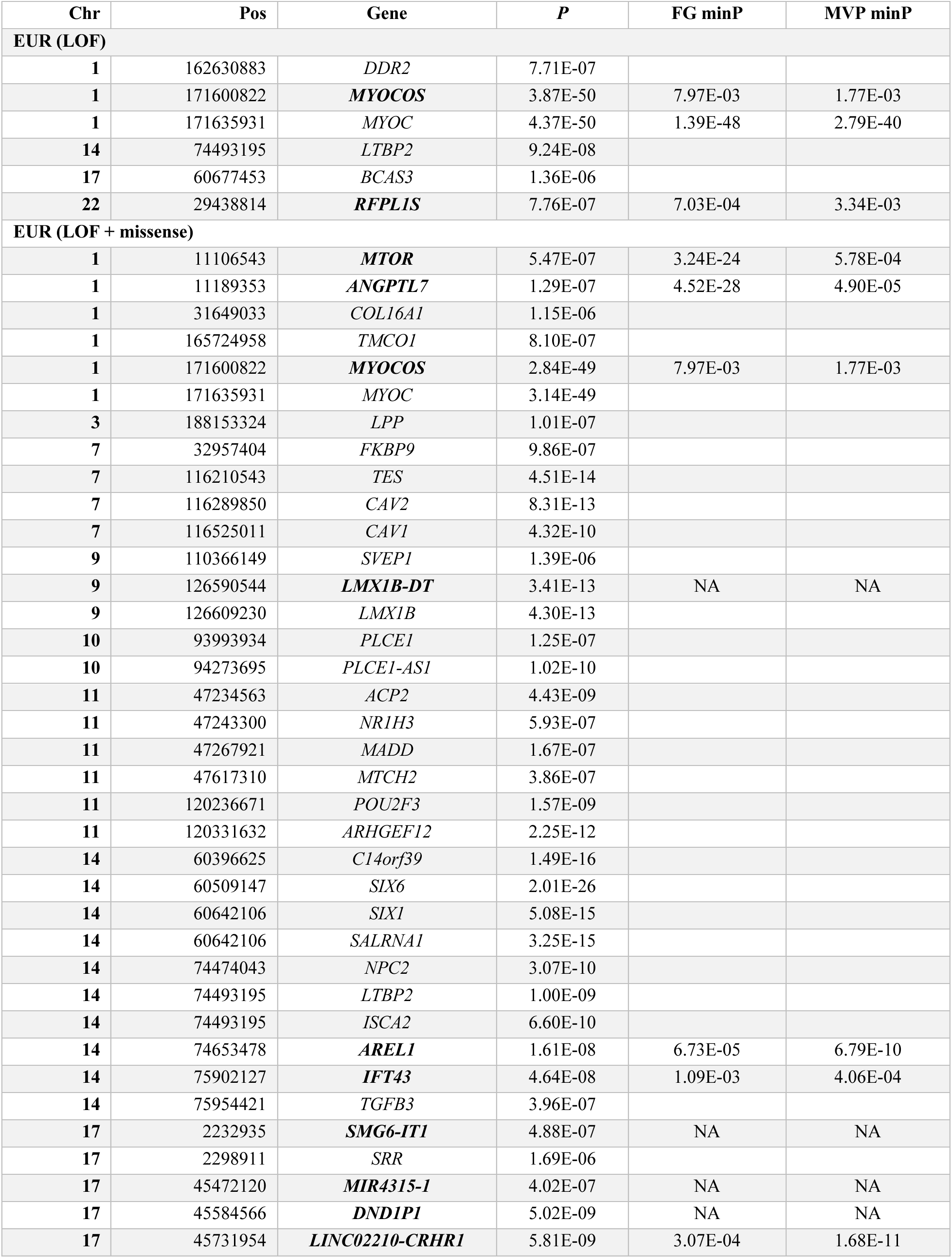
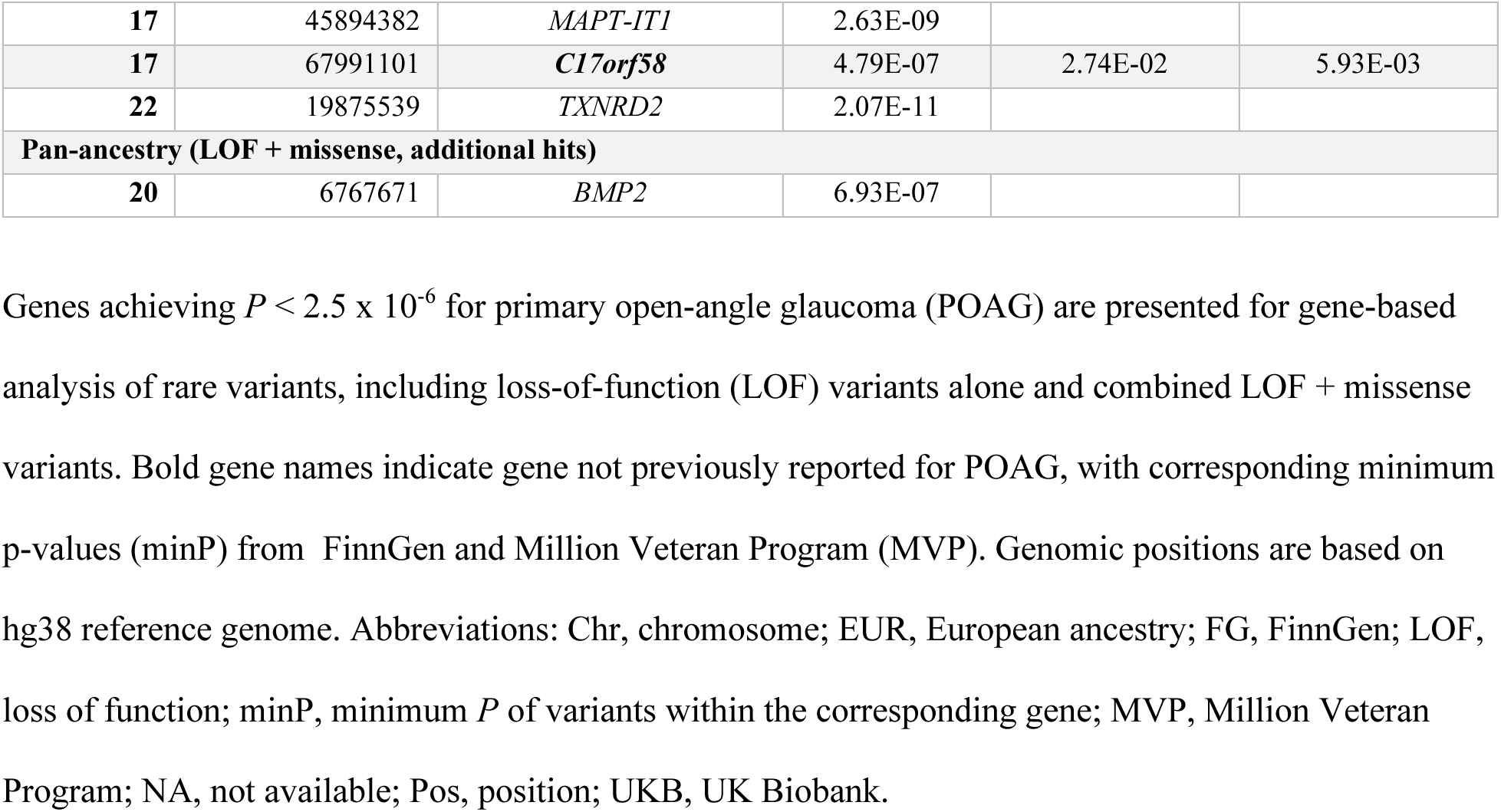
Genome-wide significant results for POAG from gene-based analysis.

Previously known POAG genes identified through common-variant studies, including *TMCO1*, *CAV2*, *CAV1*, *LMX1B*, and *SIX6*, showed significant gene-based rare-variant associations (*P* < 2.5×10^−6^).

Notably, we identified several previously unreported genes for POAG, including *MYOCOS*, *RFPL1S*, *MTOR*, *ANGPTL7*, *AREL1*, *IFT43*, *SMG6-IT1*, and *LINC02210-CRHR1*. Among these, *MTOR* and *ANGPTL7* had been previously associated with intraocular pressure (IOP)^14,15^ but not directly with POAG. Four of these novel genes, *MTOR*, *ANGPTL7*, *AREL1*, and *LINC02210-CRHR1*, showed significant associations in either FinnGen or MVP or both (minP ranging from 4.90×10^−5^ to 4.52×10^−28^). Figure 1(b) presents a circular Manhattan plot of the genome-wide p-values for gene-based pan-ancestry results.

### Gene Expression Analysis

To provide biological context for our identified previously unreported genes (68 genes, Tables 1 and 2), we evaluated their expression patterns using both bulk RNA and single-cell RNA (scRNA) datasets. We examined expression in key ocular tissues: the anterior segment drainage structures responsible for aqueous humor outflow and intraocular pressure regulation, as well as the retina and retinal ganglion cells (RGCs), the neurons that are damaged in glaucoma.

We compared our gene list to genes expressed in purified mouse Schlemm’s canal endothelial cells (SCE) identified by bulk RNA-seq^16^ (Supplementary Table 2). This bulk RNA-seq dataset represents the most comprehensive available resource for SCE, and mouse drainage structures closely resemble their human counterparts in gene expression, physiology, and morphology. Notably, 34% of our genes were expressed in SCE. Several newly identified POAG genes showed robust expression in SCE, including *ATXN7*, *COL1A2*, *DGKD*, *FAM89B*, *GPM6B*, *IFT43*, *IQGAP1*, *KBTBD4*, *LYN*, *MTOR*, *NBAS*, *NPRL2*, *OXNAD1*, *PFAS*, and *PRRC2C*. More modest SCE expression was observed for *PRRX1*, *PTEN*, *PTPRM*, *RBMS3*, *TCERG1*, *TNS3*, and *USP49*.

For trabecular meshwork (TM) gene expression analysis, we utilized scRNA-seq data from mouse and human anterior segment tissues^16–18^ (Supplementary Table 2). TM cells expressed 25% of our genes.

Several genes were enriched in TM cell clusters, including *COL1A2*, *PRRX1*, *FMO4*, *MYOCOS*, *ADGRB2*, *RBMS3*, *FAM89B*, *IFT43*, *EEF1AKNMT*, *PTPRM*, *ANGPTL7*, *TNS3*, and *GPM6B*, while *PRXL2A*, *PTEN*, and *NBAS* were present at lower levels. We identified ten genes common to both SCE and TM, as well as genes uniquely expressed in either SCE (n=12) or TM (n=7), suggesting both shared and distinct functions between these cell types.

Mining of human RGC scRNA-seq data^19^ showed that 43% of our genes were expressed in these neurons (Supplementary Table 2). RGC-enriched genes included *DPP6*, *RBMS3*, *PRRX1*, *ANKS1B*, *FMO4*, *PRXL2A*, *PLD*, *FAM89B*, *IQGAP1*, *DYDC2*, *ZNF215*, *MAT1A*, *B4GALT4-AS1*, *EEF1AKNMT*, and *PTPRM*. Additional genes (*DYDC1*, *BDH1*, *LRRC37A3*, *KBTBD4*, *NBAS*, *OXNAD1*, *ADGRB2*, *TCERG1*, *PRRC2C*, *AKAP14*, *NPRL2*, and *LRRC37A2*) showed more modest expression in RGCs. Analysis of human retina bulk RNA expression revealed that more than 90% of the identified rare-variant genes were expressed at high (55.6%) or medium (36.1%) levels (Supplementary Figure 1).

### Pleiotropy Among Genes Identified Through Rare-Variant Analyses

The identified rare-variant genes associated with POAG demonstrate significant pleiotropy, influencing a wide range of traits and conditions. Analysis using the GWAS Catalog^20,21^ highlighted that these genes (70 queried) frequently associate with various phenotypes (Supplementary Table 3), notably in categories such as cardiovascular measurements (62% of the queried genes, such as *CAV1*, *CAV2*, *LPP*, *LTBP2*, *MTCH2*, *PLCE1*, *PRRX1*, *SVEP1*, *TGFB3*, *TES*, *TXNRD2*), cancer (50% of the queried genes, such as *BCAS3*, *CAV2*, *COL1A2*, *DDR2*, *PTPRM*, *SIX1*), hematological measurements (44% of the queried genes, such as *LYN*, *PLCE1*, *MTOR*), lipid or lipoprotein measurements (38% of the queried genes, such as *FMO3*, *NR1H3*, *MTCH2*), and height (65% of the queried genes, such as *COL1A2*, *DDR2*, *MAPT-AS1*, *MTOR*, *PRRX1*, *TGFB3*). Additional notable associations include Alzheimer’s, educational attainment, refractive error, and retinal problems or measurements. Top ranked genes with the greatest number of traits associated with include *LPP* (134 mapped traits), *LINC02210-CRHR1* (91 mapped traits), and *PLCE1* (88 mapped traits). This extensive pleiotropy underscores the complex genetic landscape underlying POAG and the broad systemic impact these genes may have, potentially informing future therapeutic strategies.

### Potential Drug Targets

To identify potential therapeutic opportunities, we investigated drug targets among the identified POAG-associated genes using the Open Targets platform. Table 3 summarizes currently known drug targets for these genes, primarily identified through our gene-based analysis of rare variants. These genes are targeted for a variety of diseases and mechanisms.

**Table 3.** Known drug targets for the identified glaucoma rare-variant genes.

| Gene | Known drugs | Mechanism of action | Disease information |
| --- | --- | --- | --- |
| ***MTOR*** | Apitolisib, azd-8055, bgt-226, cc-115, dactolisib, ds-3078a, ds-7423, gedatolisib, indoximod, omipalisib, onatasertib, osi-027, palomid-529, panulisib, perhexiline, pf-04691502, pki-179, rg-7603, ridaforolimus, samotolisib, sapanisertib, sf-1126, vistusertib, voxalisib, vs-5584 | mTORC1 activator, Serine/threonine-protein kinase mTOR inhibitor | Acute myeloid leukemia, adenocarcinoma, adult glioblastoma, age-related macular degeneration, anaplastic astrocytoma, anaplastic oligodendroglioma, bladder transitional cell carcinoma, bone sarcoma, brain cancer, breast cancer, breast neoplasm, cancer, cardiovascular disease, chronic lymphocytic leukemia, clear cell renal carcinoma, colorectal cancer, colorectal neoplasm, COVID-19, Cowden disease, diabetic cardiomyopathy, diastolic heart failure, diffuse large B-cell lymphoma, endometrial cancer, endometrial carcinoma, Endometrial Clear Cell Adenocarcinoma, endometrial neoplasm, Endometrial Serous Adenocarcinoma, endometrioid carcinoma, endometrium adenocarcinoma, ependymoma, Fallopian Tube Carcinoma, gastric adenocarcinoma, glioblastoma multiforme, gliosarcoma, head and neck squamous cell carcinoma, heart failure, hepatocellular carcinoma, Hereditary breast and ovarian cancer syndrome, high grade ovarian serous adenocarcinoma, hypertrophic cardiomyopathy, idiopathic pulmonary fibrosis, infection, kidney cancer, leiomyosarcoma, leukemia, liposarcoma, lung cancer, lung carcinoma, lymphatic malformation, lymphoma, Mantle cell lymphoma, medulloblastoma, melanoma, meningioma, Merkel cell skin cancer, mesothelioma, metastasis, metastatic carcinoma, metastatic colorectal cancer, metastatic malignant neoplasm, metastatic melanoma, metastatic prostate cancer, mixed glioma, multiple myeloma, myelodysplastic syndrome, neoplasm, neoplasm of mature B-cells, neuroblastoma, neuroendocrine neoplasm, non-Hodgkins lymphoma, non-small cell lung carcinoma, osteosarcoma, ovarian cancer, ovarian clear cell cystadenocarcinoma, ovarian mucinous cystadenocarcinoma, ovarian serous cystadenocarcinoma, pancreatic adenocarcinoma, pancreatic carcinoma, pancreatic ductal adenocarcinoma, pancreatic endocrine carcinoma, pancreatic neuroendocrine tumor, pancreatic neuroendocrine tumor G1, Paraganglioma, primary myelofibrosis, primary peritoneal carcinoma, prostate cancer, renal cell adenocarcinoma, renal cell carcinoma, Richter syndrome, sarcoma, small cell lung carcinoma, soft tissue sarcoma, squamous cell carcinoma, squamous cell lung carcinoma, stomach neoplasm, T-cell acute lymphoblastic leukemia, thyroid cancer, Transitional Cell Carcinoma, triple-negative breast cancer, urinary bladder cancer, urinary bladder carcinoma, urothelial carcinoma, uterine corpus cancer, uterine neoplasm, Waldenstrom macroglobulinemia |
| ***COL1A2*** | Ocriplasmin, collagenase clostridium histolyticum | Collagen hydrolytic enzyme | Abnormal retinal morphology, Abnormality of connective tissue, contracture, decubitus ulcer, deep vein thrombosis, diabetic foot, diabetic macular edema, Dupuytren Contracture, Edema, eye disease, frozen shoulder, lipoma, macular degeneration, macular holes, Palmar Fibromatosis, Penile Fibromatosis, Peyronie disease, retinal vein occlusion, Skin ulcer, stroke, Tendinopathy, ulcer disease, Urethral stricture, uveitis |
| ***DDR2*** | Regorafenib | Discoidin domain-containing receptor 2 inhibitor | Acute myeloid leukemia, adenoid cystic carcinoma, angiosarcoma, bile duct carcinoma, biliary tract cancer, cancer, cholangiocarcinoma, colon adenocarcinoma, colorectal adenocarcinoma, colorectal cancer, colorectal carcinoma, colorectal neoplasm, cutaneous melanoma, digestive system cancer, Digestive System Carcinoma, esophageal cancer, fallopian tube cancer, gastric cancer, gastrointestinal stromal tumor, glioblastoma multiforme, grade III meningioma, hepatocellular carcinoma, intrahepatic cholangiocarcinoma, liposarcoma, liver neoplasm, macular degeneration, Malignant Bone Neoplasm, malignant colon neoplasm, malignant renal pelvis neoplasm, melanoma, metastatic colorectal cancer, myelodysplastic syndrome, neoplasm, osteosarcoma, ovarian cancer, ovarian carcinoma, ovarian neoplasm, pancreatic adenocarcinoma, pancreatic carcinoma, primary peritoneal carcinoma, rectum cancer, renal cell carcinoma, rhabdomyosarcoma, sarcoma, small cell lung carcinoma, soft tissue sarcoma, thyroid cancer |
| ***LYN*** | Bafetinib,<br>bosutinib,<br>dasatinib,<br>enmd-981693,<br>ilorasertib,<br>jnj-26483327,<br>tg100-801,<br>tolimidone, xl-228 | SRC inhibitor,<br>Tyrosine-protein<br>kinase Lyn inhibitor,<br>Tyrosine-protein<br>kinase Lyn positive<br>allosteric modulator | Acute lymphoblastic leukemia, acute myeloid leukemia, adult acute lymphoblastic leukemia, adult B acute lymphoblastic leukemia, adult glioblastoma, adult hepatocellular carcinoma, adult T acute lymphoblastic leukemia, adult T-cell leukemia/lymphoma, aging, Alzheimer disease, amyotrophic lateral sclerosis, anaplastic large cell lymphoma, angioimmunoblastic T-cell lymphoma, Autosomal dominant polycystic kidney disease, Blast Phase Chronic Myelogenous Leukemia, BCR-ABL1 Positive, brain disease, brain neoplasm, breast cancer, breast carcinoma, breast neoplasm, Burkitts lymphoma, cancer, central nervous system leukemia, Central Nervous System Neoplasm, cervical cancer, childhood acute lymphoblastic leukemia, childhood cancer, cholangiocarcinoma, choroidal neovascularization, chronic kidney disease, chronic lymphocytic leukemia, chronic myelogenous leukemia, Cognitive impairment, colorectal cancer, core binding factor acute myeloid leukemia, COVID-19, cutaneous melanoma, Dementia, diabetic retinopathy, diffuse intrinsic pontine glioma, diffuse large B-cell lymphoma, endometrial cancer, extranodal nasal NK/T cell lymphoma, fallopian tube cancer, follicular lymphoma, gastrointestinal stromal tumor, glioblastoma multiforme, gliosarcoma, head and neck malignant neoplasia, head and neck squamous cell carcinoma, hepatosplenic T-cell lymphoma, HIV infection, HIV-1 infection, Hodgkins lymphoma, idiopathic pulmonary fibrosis, leukemia, liver cancer, lung cancer, lymphoblastic lymphoma, lymphoid leukemia, lymphoma, macular degeneration, major salivary gland cancer, malignant colon neoplasm, malignant glioma, malignant pleural mesothelioma, MALT lymphoma, Mantle cell lymphoma, melanoma, mesothelioma, metastatic colorectal cancer, metastatic malignant neoplasm, metastatic melanoma, metastatic prostate cancer, multiple myeloma, mycosis fungoides, myelodysplastic syndrome, myeloid leukemia, neoplasm, neuroblastoma, nodal marginal zone B-cell lymphoma, non-Hodgkins lymphoma, non-small cell lung carcinoma, obesity, ovarian cancer, pancreatic adenocarcinoma, pancreatic carcinoma, Paraganglioma, peritoneum cancer, Pineoblastoma, polycythemia vera, prostate adenocarcinoma, prostate cancer, rectum cancer, refractory hairy cell leukemia, rhabdomyosarcoma, salivary gland adenoid cystic carcinoma, scleroderma, Sezary's disease, small cell lung carcinoma, small intestine lymphoma, Splenic Marginal Zone Lymphoma, squamous cell carcinoma, squamous cell lung carcinoma, systemic mastocytosis, T-cell large granular lymphocyte leukemia, therapy related acute myeloid leukemia and myelodysplastic syndrome, Thymoma, thymus cancer, triple-negative breast cancer, type 2 diabetes mellitus, unspecified peripheral T-cell lymphoma, urinary bladder cancer, Waldenstrom macroglobulinemia |
| ***NR1H3*** | Rovazolac,<br>bms-852927,<br>hyodeoxycholic_acid,<br>abequolixron,<br>abequolixr | Liver X receptor<br>agonist,<br>LXR-alpha modulator,<br>LXR-alpha agonist | Atopic eczema, Hypercholesterolemia, Hypercholesterolemia, endometrial cancer, non-small cell lung carcinoma |
| ***TGFB3*** | Luspatercept,<br>bintrafusp alfa,<br>fresolimumab | Transforming growth<br>factor beta inhibitor,<br>Transforming growth<br>factor beta binding<br>agent | Anemia, Aplastic anemia, Beta-thalassemia, bladder transitional cell carcinoma, brain neoplasm, breast adenocarcinoma, breast cancer, breast carcinoma, breast neoplasm, cervical adenocarcinoma, cervical cancer, cholangiocarcinoma, colorectal adenocarcinoma, diffuse scleroderma, esophageal carcinoma, focal segmental glomerulosclerosis, gallbladder cancer, gastric cancer, head and neck squamous cell carcinoma, idiopathic pulmonary fibrosis, intrahepatic cholangiocarcinoma, Kaposi's sarcoma, lung cancer, malignant pleural mesothelioma, malignant tumor of neck, mesothelioma, metastatic malignant neoplasm, metastatic prostate cancer, myelodysplastic syndrome, myeloproliferative disorder, neoplasm, non-small cell lung carcinoma, oropharynx cancer, osteogenesis imperfecta, pancreatic adenocarcinoma, pancreatic carcinoma, pancreatic neoplasm, penile cancer, prostate cancer, prostate carcinoma, renal cell carcinoma, small cell lung carcinoma, soft tissue sarcoma, Thalassemia, thymic epithelial neoplasm, Thymoma, triple-negative breast cancer, urogenital neoplasm, vaginal cancer |
| ***MAPT*** | Bepranemab,<br>gosuranemab,<br>posdinemab,<br>semorinemab,<br>tilavonemab,<br>zagotenemab | Microtubule-associated<br>protein tau inhibitor | Alzheimer disease, Classical progressive supranuclear palsy, progressive<br>supranuclear palsy |

This table shows the existing drugs that target the glaucoma rare-variant genes identified.

*MTOR* emerged as a particularly promising target, with 25 existing drugs including apitolisib, ridaforolimus, dactolisib, gedatolisib, vistusertib, and samotolisib. While primarily developed for oncology applications, *MTOR*-targeting therapeutics have expanded into cardiovascular, fibrotic, and infectious diseases. Current clinical development spans solid epithelial tumors, CNS malignancies, sarcomas, hematologic cancers, hereditary mTORopathies, and emerging non-oncological indications. *COL1A2*, targeted by ocriplasmin and collagenase clostridium histolyticum, functions as a collagen hydrolytic enzyme and is explored in conditions like diabetic macular edema, retinal vein occlusion, and macular degeneration. *NR1H3*, targeted by rovacolaz, bms-852927, hyodeoxycholic acid, abequolixron, and abequolixr, serves as a liver X receptor agonist and modulator, relevant to hypercholesterolemia and certain cancers. *LYN* is targeted by several drugs, including bafetinib, bosutinib, dasatinib, and ilorasertib, which function as SRC and tyrosine-protein kinase Lyn inhibitors primarily developed for acute leukemias and chronic myeloid leukemia. *DDR2* is targeted by regorafenib, a discoidin domain-containing receptor 2 inhibitor currently being evaluated for acute myeloid leukemia and macular degeneration. *TGFB3*-targeting agents, including luspatercept, bintrafusp alfa, and fresolimumab, inhibit transforming growth factor-beta signaling and are being investigated across solid tumors, bone-marrow failure syndromes, and fibrotic conditions. *MAPT* is targeted by tau inhibitors such as bepranemab, gosuranemab, and semorinemab, which are being actively studied for Alzheimer’s disease and related tauopathies.

Notably, among these genes, *MTOR*, *COL1A2*, *LYN*, and *DDR2* have therapeutic investigations relevant to macular degeneration, while *LYN* and *MAPT* are associated with treatments targeting Alzheimer’s disease, suggesting potential mechanistic overlap with POAG pathophysiology.

## Discussion

In this study, we conducted the largest WGS analysis of glaucoma to date using data from UKB. By employing complementary single-variant and gene-based analytical frameworks, we have significantly expanded our understanding of the genetic architecture of POAG, particularly regarding the contribution of rare variants that were largely missed in previous microarray-based studies. In addition to confirming known POAG-associated genes, we identified 68 previously unreported genes, demonstrating the enhanced discovery power of WGS data. Approximately half of these novel genes were also associated with glaucoma in independent cohorts from FinnGen, MVP, or both, providing crucial validation. Notably, seven of these genes represent drug targets for existing clinical treatments, highlighting immediate translational potential.

WGS enabled the identification of rare-variant associations with POAG through both single-variant and gene-based analyses. Our single-variant analysis revealed many novel rare variants significantly associated with POAG, including rs147235365 in *FMO3*, rs534598623 in *PRRC2C*, rs142017159 in *PRRX1*, rs745475536 in *FMO4*, and rs756856153 in *EEF1AKNMT*. Through gene-based analysis, we identified 45 genes harboring rare variants with significant POAG associations. Six of these genes were identified from LOF variants alone, while the majority (87%) were identified from a combination of LOF and missense variants, highlighting the importance of including missense variants in rare-variant analysis. Among the 45 genes, 12 of these genes were previously unreported for POAG (though *ANGPTL7* and *MTOR* had been associated with intraocular pressure) and 73% overlapped with previously reported genes for POAG identified through array-based GWASs, reaffirming the importance of these genes. Though replication remains challenging as most existing GWASs rely on arrays and imputation, our study provides the first evidence of rare-variant associations with POAG for these genes.

Beyond rare variants, WGS also identified numerous common variants and genes missed by previous large-scale GWAS meta-analyses. These include common variants in *MIR3125*, *MAT1A*, *DYDC1*, *DYDC2*, *PRXL2A*, *AREL1*, *IQGAP1*, *NSFP1*, *SEPTIN9*, and *WNT7B*. These associations were replicated in FinnGen, MVP, or both cohorts (*P* < 5×10^−5^), demonstrating that a substantial number of common variants and genes remain undetected by array-based approaches and imputation.

There has been growing interest in pleiotropy within human genetics studies. Through GWASs, thousands of genetic associations have been identified, and investigating whether traits share common genetic variants can illuminate underlying biological processes and reveal important therapeutic opportunities, including drug repositioning strategies^22,23^. Our study demonstrates that rare-variant genes associated with POAG exhibit extensive pleiotropy with both ocular and systemic traits. Among ocular phenotypes, several of our identified genes are also associated with refractive error, retinal degeneration, and various retinal measurements, suggesting shared pathways in ocular development and pathology. More broadly, numerous systemic traits were associated with our identified POAG genes, including Alzheimer’s disease, cardiovascular measurements, cancer, hematological parameters, and lipid or lipoprotein levels. This pleiotropic pattern suggests that POAG shares genetic architecture with multiple disease processes, potentially explaining epidemiological associations observed between glaucoma and systemic conditions. From a therapeutic perspective, pleiotropy indicates that drugs developed for one condition may have efficacy in treating related disorders. Notably, about 10% of our rare-variant genes, including *MTOR*, *COL1A2*, *DDR2*, *LYN*, *NR1H3*, *TGFB3*, and *MAPT*, are established drug targets with existing therapeutic compounds. This finding highlights immediate opportunities for drug repositioning in glaucoma treatment. Furthermore, the remaining genes in our dataset represent potential novel therapeutic targets that warrant further investigation for drug development efforts.

While our analysis participants were primarily of EUR ancestry, we demonstrated that including subjects of other ancestries in a pan-ancestry analysis further enhanced statistical power. The inclusion of non-EUR participants, despite comprising only 5% of the UKB cohort, enabled the identification of 14 additional previously unreported genes for POAG. This underscores the importance of diverse ancestral representation in genetic studies and suggests that as more genetic data from diverse populations become available, we can anticipate further novel discoveries.

This study has several limitations. Despite being the largest WGS dataset currently available, ancestral diversity remains low, with participants of EUR ancestry comprising approximately 95% of the UKB cohort. Larger, more diverse datasets, such as those being developed in the All of Us project, will be invaluable for further discovery and validation. Rare variants present intrinsic challenges due to their low frequency, making replication substantially more difficult than for common variants. Most rare variants identified in this study lack exact counterparts in FinnGen and MVP GWAS summary statistics. Additionally, our use of combined self-reported glaucoma and ICD-10/9 codes for phenotyping in UKB may lack homogeneity for specialized glaucoma subtypes, but given POAG accounts for the majority of glaucoma and previous studies have validated this approach for studying POAG genetics^24,25^. Furthermore, our analysis did not cover intergenic variants, which are functionally challenging to interpret.

In conclusion, we have conducted the largest WGS study of POAG to date, demonstrating the efficacy of both single-variant and gene-based analyses. Our results strongly support the value of gene-based aggregation and pan-ancestry approaches for increasing study power, particularly for rare variant discovery. This study highlights the substantial value of WGS in enhancing our understanding of the biological mechanisms underlying this blinding disease and has uncovered potential novel therapeutic targets for glaucoma treatment.

## Materials and Methods

### UKB Resource

The UK Biobank (UKB) is an ongoing large-scale prospective cohort study, the details of which have been described extensively elsewhere^26,27^. Briefly, UKB recruited over 500,000 adult participants aged 40-70 years at enrollment who were living in the United Kingdom and registered with the National Health Service at study baseline (2006-2010). Comprehensive data collection included medical information from both self-reports and electronic health records, family history, lifestyle information, and DNA samples.

Glaucoma cases were identified using a combination of self-reported glaucoma (UKB data fields 6148, 20002) and ICD-10 or ICD-9 diagnosis codes for glaucoma (UKB data fields 131186, 131188, 41202, 41204, 41076, 41078, 41270). We excluded cases of secondary glaucoma resulting from eye trauma, inflammation, other eye disorders, drug-induced effects, or other specific glaucoma types. This approach for glaucoma case identification based on self-reports and ICD codes has been validated in previous studies^24,25^, and the proportion of non-primary open-angle glaucoma (POAG) cases in UKB is expected to be minimal^28^. Controls were defined as participants without glaucoma or self-reported eye problems. Our access to this resource was approved by UKB (application number 23424), and we obtained access to fully de-identified data.

Whole-genome sequencing of all UKB participants was performed using the Illumina NovaSeq 6000 platform to an average coverage of 32.5×. Detailed protocols for sequencing, variant calling, and quality control have been described previously^29^. High-quality variants comprised over 98.8% of single nucleotide polymorphisms (SNPs) and 96.05% of indels (AA score > 0.5 and < 5 duplicate inconsistencies)^29^. We utilized SNPs and indels called using GraphTyper and retained samples with missing rates < 1%. Variants in gene regions (excluding intergenic ones) with call rates > 95% and minor allele count (MAC) > 1 were included. Variant annotation was performed using VEP^2^ and ANNOVAR^30^. After overlapping with phenotype and covariate data, the final dataset comprised 18,171 EUR cases and 446,607 EUR controls, and 19,440 pan-ancestry cases and 469,448 pan-ancestry controls.

### Single-variant and Gene-based Analyses

Single-variant association analyses were performed using REGENIE^13^, a machine-learning method that accounts for population stratification and sample relatedness. We analyzed all non-intergenic variants with MAC ≥ 5 (REGENIE default threshold) and included age, sex, and the first 10 principal components (PCs) of genetic ancestry as covariates. Variants achieving *P* < 5 × 10^−8^ were considered genome-wide significant, consistent with established GWAS standards. We performed both EUR-only and pan-ancestry analyses. For pan-ancestry analyses, we included an additional covariate representing five major ancestral groups (European, South Asian, East Asian, African, and Admixed American) identified by projecting PCs onto the 1000 Genomes Project reference panel.

Gene-based association tests were also conducted using REGENIE^13^, which implements rare-variant collapsing and aggregation tests including SKAT-O^31^, burden^32^, and SKAT^33,34^ methods. We constructed gene sets using predicted loss-of-function (LOF) variants, defined as stop gained, stop lost, start lost, splice donor, splice acceptor, and frameshift variants based on VEP^35^ annotation and gnomAD LOF classification^36^. Additionally, we analyzed gene sets combining LOF and missense variants. Age, sex, and the first 10 PCs were included as covariates. Genes achieving *P* < 2.5 × 10^−6^ were considered significant. Both EUR-only and pan-ancestry analyses were performed, with dummy variables for major ancestral groups added as additional covariates in the latter.

### Glaucoma Lookup in FinnGen and MVP

For replication of our findings, we performed lookups of all genome-wide significant POAG variants and genes in publicly available summary statistics from the FinnGen and MVP resources.

FinnGen is a large-scale biobank study focused on the Finnish population^37^. The study has enrolled over 500,000 participants who have been genotyped and phenotyped. The study leverages the power of nationwide biobanks, electronic health records, and population isolation to identify variants associated with various diseases. Data collection from Finnish biobanks and digital healthcare records began in 2017, with a recruited population averaging 63 years of age and predominantly hospital-based recruitment. Phenotypes were constructed using ICD-9 and ICD-10 codes. Genotyping was performed using custom Axiom FinnGen1 and legacy arrays, followed by imputation to 21.3 million markers based on whole-genome sequences from Finnish individuals. We utilized the DF12 release of their POAG association results, which included 10,832 cases and 473,757 controls, analyzed using SAIGE mixed models.

The Department of Veterans Affairs Million Veteran Program (MVP) is an ongoing prospective cohort study and mega-biobank established in 2011 to investigate genetic influences on health and disease among veterans^38^. The program reached its milestone of one million enrolled participants in 2023. The study population is ancestrally diverse, including participants of European, African, Admixed American, and East Asian ancestry, with a mean age of 61.9 years and 91.2% male composition. Genotyping was performed using a custom ThermoFisher Axiom MVP 1.0 platform, followed by imputation to a hybrid reference panel comprising the African Genome Resources panel and 1000 Genomes Project (Phase 3 v5). We utilized the recently released POAG association results for European ancestry participants, which included 20,401 cases and 431,032 controls, analyzed using SAIGE mixed models.

### Gene Expression

We utilized Genevestigator^39^, a web-based gene expression database, to query bulk RNA information in different human tissues, including retina. RNA information for Schlemm’s canal endothelium (SCE) were generated from mouse bulk RNA-seq data^16^. Single-cell expression data were curated from mouse and human TM cells using the Single Cell Portal and CellxGene resources, based on published datasets^16–18^. Single-cell expression data for RGC were obtained from previously published results^19^.

### GWAS Catalog Gene Lookup

For checking broader phenotypic associations of the rare-variant genes, we used the GWAS Catalog (May 2025 version), a database of all published GWASs maintained by NHGRI and EBML-EBI^20,21^. The GWAS Catalog provides an excellent resource for studying pleiotropy in humans^40^.

### Drug Targets Prioritization

To prioritize drug targets for the identified rare-variant genes, we used the Open Targets Platform^41^. For the identified genes, we queried the Open Targets for known drugs, their mechanisms of action (source ChEMBL), and disease information. The druggable genes provide key information on the relevance of these genes and potential drugs for repurposing for glaucoma management.

## Data availability

Access to UK Biobank data, both phenotypic and genetic, is available to bona fide researchers through application on the UK Biobank website (https://www.ukbiobank.ac.uk). Our use of UK Biobank data was performed under application number 23424.

The URLs for accessed databases:

ChEMBL, https://www.ebi.ac.uk/chembl/

FinnGen, https://www.finngen.fi/

Genevestigator, https://genevestigator.com/

MVP, https://www.ncbi.nlm.nih.gov/projects/gap/

Open Target Platform, https://platform.opentargets.org

UK Biobank, https://www.ukbiobank.ac.uk

## Code availability

The programs used for data analysis:

ANNOVAR, http://annovar.openbioinformatics.org/

FinnGen, https://www.finngen.fi/

REGENIE, https://rgcgithub.github.io/regenie/

VEP, https://useast.ensembl.org/info/docs/tools/vep/index.html

R, https://www.r-project.org

## Acknowledgements

We would like to thank the study participants and investigators from the UK Biobank as well as the staff who aided in data collection and processing. We also want to acknowledge the participants and investigators of the FinnGen study and the Million Veteran Program.

## Author contributions

X.R.G. conceived, planned and oversaw the present study. X.R.G. analyzed the data. K.K. queried gene expression databases. X.R.G. and K.K. wrote the manuscript.

